# Deep learning driven quantification of interstitial fibrosis in kidney biopsies

**DOI:** 10.1101/2021.01.03.21249179

**Authors:** Yi Zheng, Clarissa A. Cassol, Saemi Jung, Divya Veerapaneni, Vipul C. Chitalia, Kevin Ren, Shubha S. Bellur, Peter Boor, Laura M. Barisoni, Sushrut S. Waikar, Margrit Betke, Vijaya B. Kolachalama

## Abstract

Interstitial fibrosis and tubular atrophy (IFTA) on a renal biopsy are strong indicators of disease chronicity and prognosis. Techniques that are typically used for IFTA grading remain manual, leading to variability among pathologists. Accurate IFTA estimation using computational techniques can reduce this variability and provide quantitative assessment by capturing the pathologic features. Using trichrome-stained whole slide images (WSIs) processed from human renal biopsies, we developed a deep learning (DL) framework that captured finer pathological structures at high resolution and overall context at the WSI-level to predict IFTA grade. WSIs (n=67) were obtained from The Ohio State University Wexner Medical Center (OSUWMC). Five nephropathologists independently reviewed them and provided fibrosis scores that were converted to IFTA grades: <=10% (None or minimal), 11-25% (Mild), 26-50% (Moderate), and >50% (Severe). The model was developed by associating the WSIs with the IFTA grade determined by majority voting (reference estimate). Model performance was evaluated on WSIs (n=28) obtained from the Kidney Precision Medicine Project (KPMP). There was good agreement on the IFTA grading between the pathologists and the reference estimate (Kappa=0.622±0.071). The accuracy of the DL model was 71.8±5.3% on OSUWMC and 65.0±4.2% on KPMP datasets, respectively. Identification of salient image regions by combining microscopic and WSI-level pathological features yielded visual representations that were consistent with the pathologist-based IFTA grading. Our approach to analyzing microscopic- and WSI-level changes in renal biopsies attempts to mimic the pathologist and provides a regional and contextual estimation of IFTA. Such methods can assist clinicopathologic diagnosis.

**Translational statement:** Pathologists rely on interstitial fibrosis and tubular atrophy (IFTA) to indicate chronicity in kidney biopsies and provide a prognostic indicator of renal survival. Although guidelines for evaluation of IFTA exist, there is variability in IFTA estimation among pathologists. In this work, digitized kidney biopsies were independently reviewed by five nephropathologists and majority voting on their ratings was used to determine the IFTA grade. Using this information, a deep learning model was developed that captured microscopic and holistic features on the digitized biopsies and accurately predicted the IFTA grade. The study illustrates that deep learning can be utilized effectively to perform IFTA grading, thus enhancing conventional clinicopathologic diagnosis.

## Introduction

Renal biopsy is an integral part of clinical work-up for patients with several kidney diseases (1), as it provides diagnostic and prognostic information that guides treatment. Despite such integral clinical use, current assessment of renal biopsy suffers from some limitations (2). Evaluation of clinically relevant pathological features such as the amount of interstitial fibrosis and tubular atrophy (IFTA), an important prognostic indicator, is based mainly on visual estimation and semi-quantitative grading and hence cannot reveal relationships that are not immediately evident using compartmentalized approaches (3). Moreover, renal biopsies typically contain other elements (e.g. glomeruli, arteries and arterioles, etc.) that are interspersed throughout the renal parenchyma and preclude an accurate IFTA assessment by the human eye. Such estimates do not capture finer details or heterogeneity across an entire slide, and therefore may not be optimal for analyzing renal tissues with complex histopathology. These aspects underscore the need for leveraging advances in digital pathology and developing modern data analytic technologies such as deep learning (DL) for comprehensive image analysis of kidney pathology.

DL techniques that utilize digitized images of biopsies are increasingly considered to facilitate the routine workflow of a pathologist. There has been a surge of publications showcasing DL applications in clinical medicine and biomedical research, with a few of them in nephrology and nephropathology (4-9). Specifically, DL techniques such as convolutional neural networks have been widely used for the analysis of histopathological images. In the context of kidney diseases, researchers have been able to produce highly accurate methods to evaluate disease grade, segment various kidney structures, as well as predict clinical phenotypes (10-18). While this body of work is highly valuable, almost all of it focuses on analyzing high-resolution whole slide images (WSIs) by breaking them down into smaller patches (or tiles) or resizing the images to a lower resolution, and associating them with various outputs of interest. These techniques have various advantages and limitations. While the patch-based approaches maintain image resolution, analyzing each patch independently cannot preserve the spatial relevance of that patch in the context of the entire WSI. In contrast, resizing the WSI to a lower resolution can be a computationally efficient approach but may not allow one to capture the finer details present within a high-resolution WSI.

The goal of this study was to develop a computational pipeline that can process WSIs to accurately capture the IFTA grade. To achieve this goal, we attempted to emulate the nephropathologist’s approach to grading the biopsy slides under a microscope. A typical workflow by the expert involves manual operations such as panning as well as zooming in and out of specific regions on the slide to evaluate various aspects of the pathology. In the ‘zoom out’ assessment, pathologists review the entire slide and perform ‘global’ or ‘WSI-level’ evaluation of the kidney core. In the ‘zoom in’ assessment, they perform in-depth, microscopic evaluation of ‘local’ pathology in the regions of interest. Both these assessments allow them to comprehensively assess the kidney biopsy, including estimation of IFTA grade. We hypothesized that a computational approach based on DL would mimic the process that nephopathologists use when evaluating the kidney biopsy images. Using WSIs and their corresponding IFTA grades from two distinct cohorts, we addressed the following objectives. First, the framework needs to process image sub-regions (or patches) and quantify the extent of IFTA within those patches. Second, the framework needs to process each image patch in the context of its environment and assess IFTA on the WSI. We developed a computational pipeline based on deep learning that can incorporate patterns and features from the local patches along with information from the WSI in its entirety to provide context for the patches. Through this combination of patch-level and global-level data, the model was designed to accurately predict IFTA grade. We brought together an international team of practicing nephropathologists who evaluated the digitized biopsies and provided the IFTA grades. The WSIs and their corresponding IFTA grades were used to train and validate the deep learning model. We also compared the deep learning model with a modeling framework based on traditional computer vision and machine learning that uses image descriptors and textural features. We then reported the performances of the deep learning model and identified image sub-regions that are highly associated with the IFTA grade.

## Methods

### Study population, slide digitization and pre-processing

We obtained digitized images of trichrome-stained kidney biopsies of patients submitted to The Ohio State University Wexner Medical Center (OSUWMC) and obtained WSIs from the Kidney Precision Medicine Project (KPMP) (19), which is a multi-year project funded by the NIDDK with the purpose of understanding and finding new ways to treat chronic kidney disease and acute kidney injury. Renal biopsy as well as patient data collection, staining and digitization followed protocols approved by the Institutional Review Board at OSUWMC (Study number: 2018H0495) (**Table 1A**). The WSIs were de-identified and uploaded to a secure, web-based software (PixelView, deepPath, Inc.). C.A.C. served as the group administrator of the software account and provided separate access to the WSIs to the other nephropathologists (K.R., S.S.B., L.M.B. and P.B.), who were assigned as users to the group account. This process allowed each expert to independently evaluate the digitized biopsies. The WSIs and relevant data from KPMP were obtained following review and approval of the Data Usage Agreement between KPMP and Boston University (**Tables 1B & S1**). All methods were performed according to federal guidelines and regulations. Renal tissues consisted of needle biopsy samples from biopsies received at OSUWMC and KPMP participants. All OSUWMC biopsies were scanned using a whole slide image (WSI) scanner (Aperio, Leica Biosystems or NanoZoomer, Hamamatsu) at 40x apparent magnification, resulting in WSIs with resolution 0.25 µm per pixel (**Figure S1**). All the WSIs from KPMP were generated by digitizing renal biopsies using Aperio AT2 high volume scanners (Leica Biosystems) at 40x apparent magnification with resolution 0.25 µm per pixel (**Figure S1**). More details on the pathology protocol can be obtained directly from the KPMP website (https://www.kpmp.org). The Aperio-based WSIs were obtained in the SVS format and the Hamamatsu-based WSIs were obtained in NDPI format, respectively.

**Table 1.**
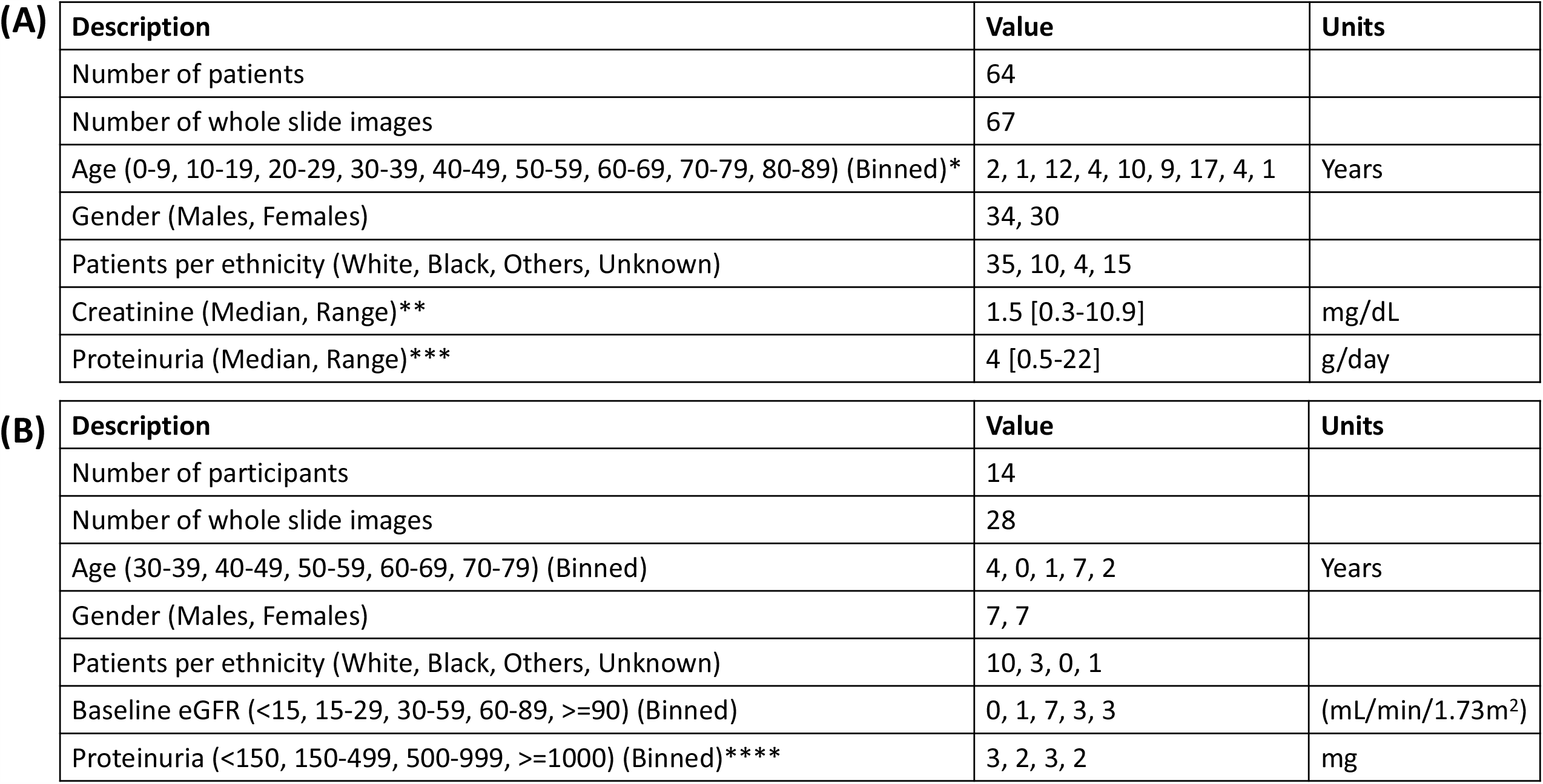
Study population. (A) The cases obtained from Ohio State University Wexner Medical Center are shown. A single trichrome stained biopsy slide was digitized for each patient. *Age was unavailable on 4 patients. **Creatinine values were unavailable on 11 patients. ***Proteinuria values were unavailable on 13 patients. Baseline eGFR data was unavailable on all the patients. (B) All the cases obtained from the Kidney Precision Medicine Project (KPMP) are shown. For each participant, two trichrome images were available. Creatinine values were unavailable on all the participants. ****Proteinuria values were unavailable on 4 participants.

| (A) | Description | Value | Units |
| --- | --- | --- | --- |
|  | Number of patients | 64 |  |
|  | Number of whole slide images | 67 |  |
|  | Age (0-9, 10-19, 20-29, 30-39, 40-49, 50-59, 60-69, 70-79, 80-89) (Binned)* | 2, 1, 12, 4, 10, 9, 17, 4, 1 | Years |
|  | Gender (Males, Females) | 34, 30 |  |
|  | Patients per ethnicity (White, Black, Others, Unknown) | 35, 10, 4, 15 |  |
|  | Creatinine (Median, Range)** | 1.5 [0.3-10.9] | mg/dL |
|  | Proteinuria (Median, Range)*** | 4 [0.5-22] | g/day |

| (B) | Description | Value | Units |
| --- | --- | --- | --- |
|  | Number of participants | 14 |  |
|  | Number of whole slide images | 28 |  |
|  | Age (30-39, 40-49, 50-59, 60-69, 70-79) (Binned) | 4, 0, 1, 7, 2 | Years |
|  | Gender (Males, Females) | 7, 7 |  |
|  | Patients per ethnicity (White, Black, Others, Unknown) | 10, 3, 0, 1 |  |
|  | Baseline eGFR (<15, 15-29, 30-59, 60-89, >=90) (Binned) | 0, 1, 7, 3, 3 | (mL/min/1.73m²) |
|  | Proteinuria (<150, 150-499, 500-999, >=1000) (Binned)**** | 3, 2, 3, 2 | mg |

A manual quality check was performed on all the WSIs by a nephropathologist (C.A.C.). This check ensured there were no artifacts on the selected WSI regions such as air bubbles, folding, compressing, tearing, over- or under-staining, stain batch variations, knife chatter and thickness variances. Since most WSIs had multiple cores, the nephropathologist was able to select a core that had no quality issues on all cases (**Figure S2**). The selected portion of the WSI was then carefully cropped and converted to numeric matrices for further analysis.

### Fibrosis grading

A nephropathologist (C.A.C.) identified and annotated the cortical regions within each WSI (**Figure 1**) where both cortex and medulla were present. All the nephropathologists accessed and independently reviewed the WSIs for IFTA using a web browser on their respective computers. The score was provided as percentage of cortical regions with IFTA (0-100%), which was then converted to a semi-quantitative grade: <=10% (None or minimal), 11-25% (Mild), 26-50% (Moderate), and >50% (Severe) (20). The final IFTA grades were computed by performing majority voting on the grades obtained from each nephropathologist. The fibrosis scores for the KPMP dataset were obtained directly from the study investigators and converted to IFTA grades using the same criterion. The derived IFTA grades from both datasets were used for further analysis.

**Figure 1.**
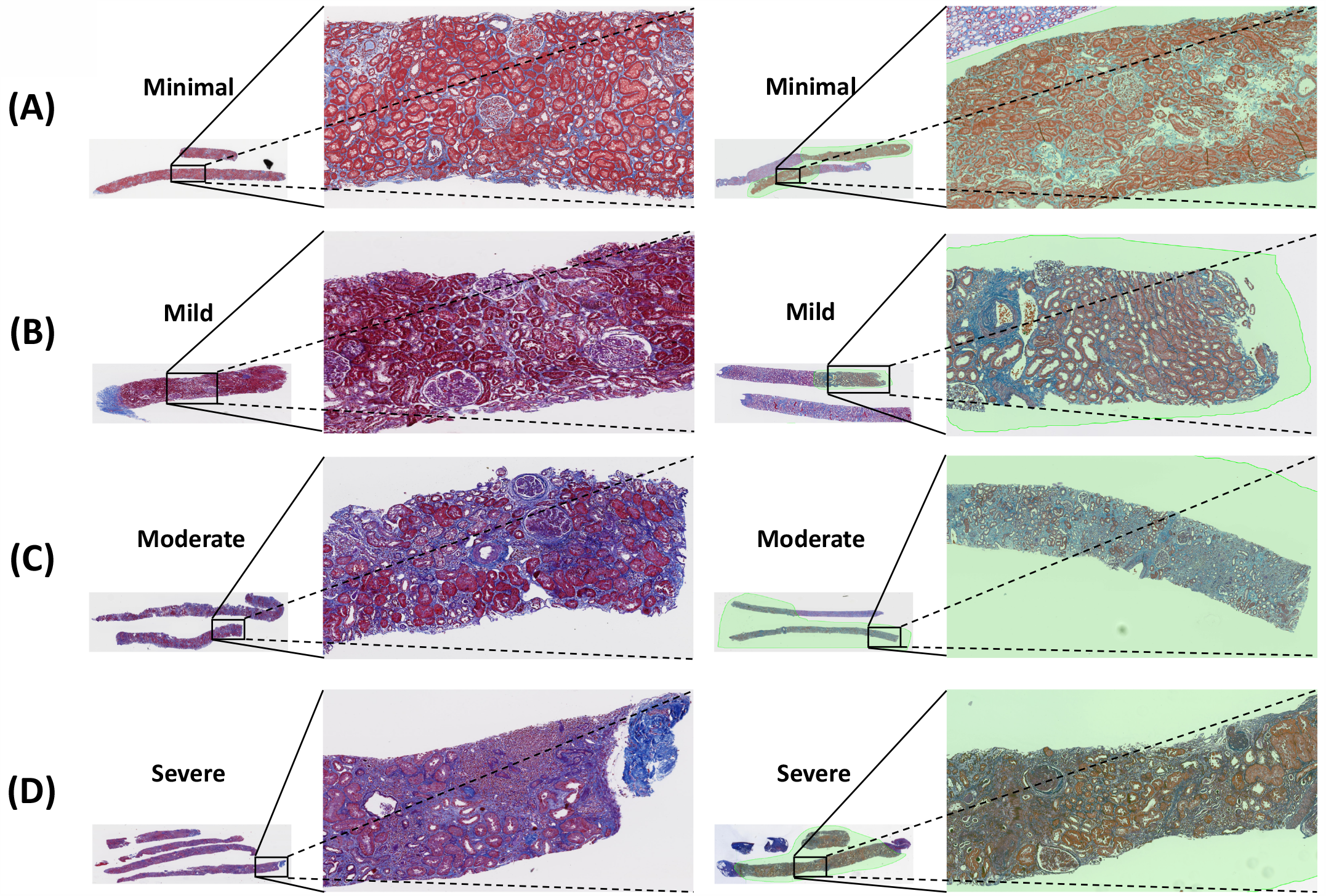
Trichrome stained whole slide images of human renal biopsies. Sample trichrome images are shown on cases graded as (A) Minimal IFTA, (B) Mild IFTA, (C) Moderate IFTA and (D) Severe IFTA. For each grade, two different images are shown, where the left image had no annotations because the entire image comprised of the cortical region. On the images on the right side, a nephropathologist (C.A.C.) annotated the cortical regions. For cases with no annotations, the entire image served as inputs to the DL model, and for cases with annotations, the annotated regions were segmented, which served as inputs to the DL model. The final IFTA grading was derived by performing majority voting on the ratings obtained from five nephropathologists.

### Deep learning framework

Our DL architecture is based on combining the features learned at the global level of the WSI along with the ones learned from local high-resolution image patches from the WSI (**Figure 2A**). Similar DL architectures have been recently applied on a few computer vision-related tasks (21-25). In what follows, we refer to this architecture as *glpathnet*. Briefly, glpathnet comprises three arms: (a) local branch (**Figure 2A** top), (b) global branch (**Figure 2A** bottom) and (c) ensemble branch (**Figure 2A** right). The local branch receives cropped filtered patches from the original images as the input to a Feature Pyramid Network (FPN) model (26). The FPN uses an efficient architecture to leverage multiple feature maps at low and high resolutions to detect objects at different scales.

**Figure 2.**
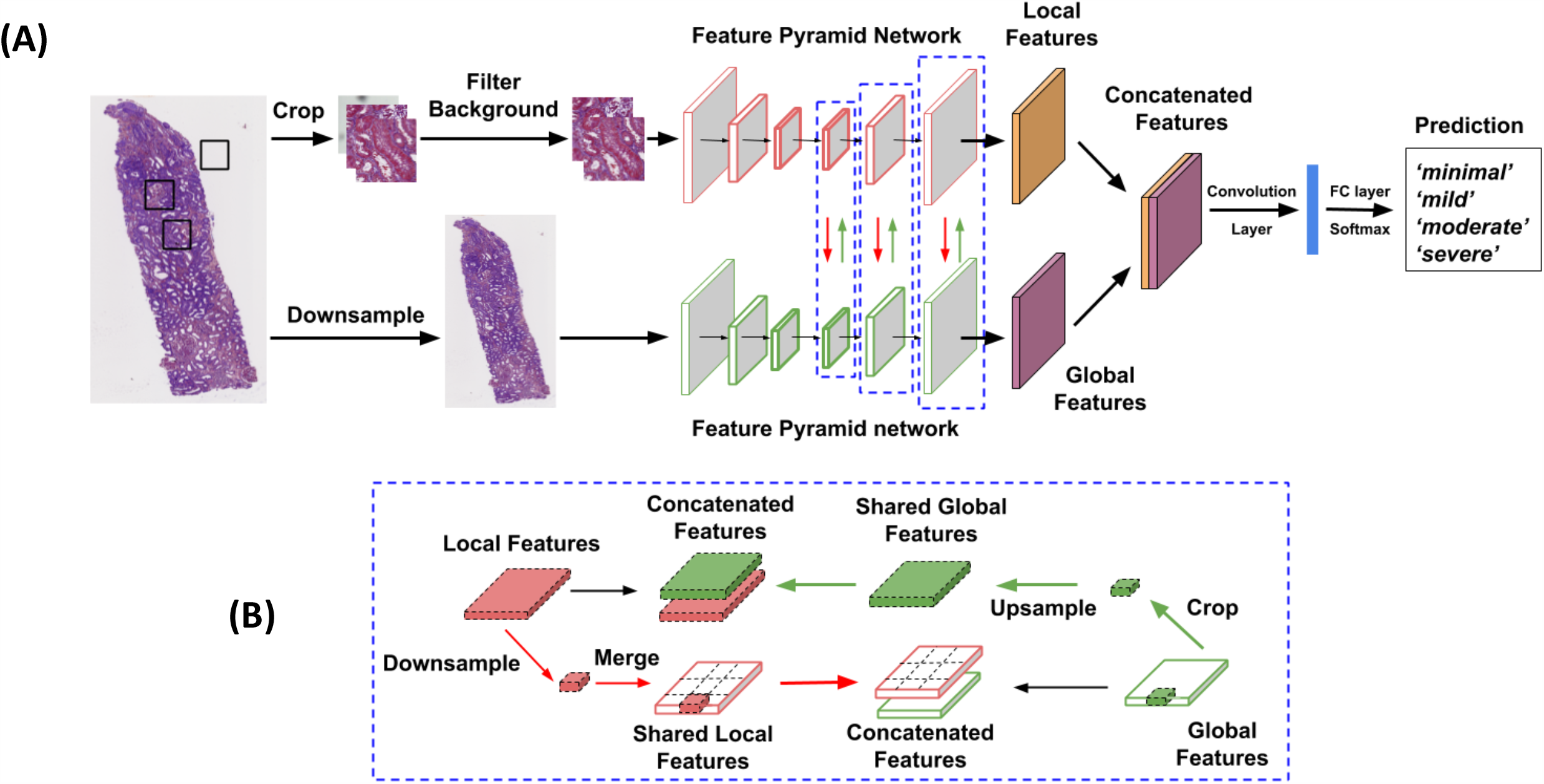
Deep learning architecture. (A) The proposed deep neural network uses a novel approach that learns from both local and global image features to predict the output label of interest. The local features are learned at the level of image patches and the global features are learned on a downsampled version of the whole image. (B) A schematic showing local and global feature sharing is shown. The current local and global feature maps that are fused at each layer, where each layer is represented as a blue-dashed box.

Cropped image patches (N_p_ x N_p_ pixels) were automatically extracted from the original WSIs and labelled as “tissue” or “background” using the following criterion. Image patches containing tissue within at least 50% or more pixels were labeled as “tissue” and labeled as “background” otherwise. The local branch containing the image patches labeled as “tissue” are fed into the FPN model. The global branch contains down-sampled low-resolution versions (N_g_ x N_g_ pixels) of the original WSIs, which serve as inputs to another FPN model. To enable local and global feature interaction, the feature maps from all layers of either branches are shared with the others (**Figure 2B**). The global branch crops its feature maps at the same spatial location of the current local patch. To interact with the local branch, glpathnet upsamples the cropped feature maps to the same size of the feature maps from the local branch in the layer with the same depth. Subsequently, glpathnet concatenates the local feature maps and cropped global feature maps, which are fed into the next layer in the local branch. In a symmetrical fashion, the local branch down-samples its feature maps to the same relative spatial ratio as the patches cropped from the original input image. Based on the location of the cropped patches, the down-sampled local feature maps are merged together into feature maps of the same size of the global branch feature in the same layer. For the patches labeled as “background”, we used feature maps with all zeros. The global feature maps and merged local feature maps are concatenated and fed into the next layer in the global branch.

The ensemble branch in glpathnet contains a convolutional layer followed by a fully-connected layer. It takes the concatenated feature maps from the last layer of the local branch and the same ones from the global branch. The output of the ensemble branch is a patch-level IFTA grade and the final IFTA grade was determined as the most common patch-level IFTA grade.

We used cross-entropy loss to train glpathnet on the OSUWMC data using a pre-trained DL architecture (ResNet50) (27), as part of the convolutional network of the FPN model. In order to maximize efficiency, both N_p_ and N_g_ were set to 508 pixels. We used ADAM optimizer (*β*_1_=0.9, *β*_2_=0.999) to optimize model training with a batch size of 6. We assigned the initial learning rate to 2×10^−5^ for the local branch, and 1×10^−4^ for the global branch, respectively. We implemented glpathnet using PyTorch and model training was performed on a GPU workstation containing NVIDIA’s GeForce RTX 2080 Ti graphics cards with a 11-Gb GDDR6 memory. Model training took less than 16 hours to reach convergence. Prediction of IFTA grade on a new WSI that was not used for model training took approximately 30 seconds.

### Traditional machine learning model

For comparison, we constructed an IFTA classification model based on traditional machine learning that used derived features from OSUWMC WSI data. We used *Weighted Neighbor Distance using Compound Hierarchy of Algorithms Representing Morphology* (WND-CHARM) (28-30), which is a multi-purpose image classifier that can extract ∼3,000 generic image descriptors including polynomial decompositions, high contrast features, pixel statistics, and textures (**Table S2**). These features were directly derived from the raw WSI, transforms of the WSI, and compound transforms of the WSI (transforms of transforms). Using these features as inputs, we constructed a 4-label classifier to predict the final IFTA grade. Model was trained on the OSUWMC dataset and the KPMP dataset was used for testing.

### Performance metrics

The final IFTA grade (reference estimate) was determined by taking the majority vote on the IFTA grading among all the five nephropathologists. The agreement between the nephropathologists was computed using Kappa (*κ*) scores between each pathologist grade and the reference estimate. The percentage agreement between the pathologists and between pathologists and the reference estimate was also computed. For the DL model trained on the OSUWMC dataset, a 5-fold cross validation was performed and the average model accuracy, sensitivity/recall, specificity, precision and kappa scores were reported. Sensitivity/recall measures the proportion of true positives that are correctly identified, specificity measures the proportion of true negatives that are correctly identified, and precision is a fraction of true positives over the total number of positive calls.

### Data sharing

Data from the OSUWMC can be obtained upon request and subject to institutional approval. Data from KPMP can be freely downloaded from https://www.kpmp.org.

## Results

There was good agreement on the IFTA grading between the nephropathologists, where pairwise agreements ranged from 0.48 to 0.63 (**Figure 3A**). Inter-pathologist ratings assessed using pairwise Kappa (*κ*) showed moderate agreement, ranging from 0.31 to 0.50 (**Figure 3B**). There was good agreement when each pathologist grading was compared with the reference IFTA grade (*κ*=0.622±0.071).

**Figure 3.**
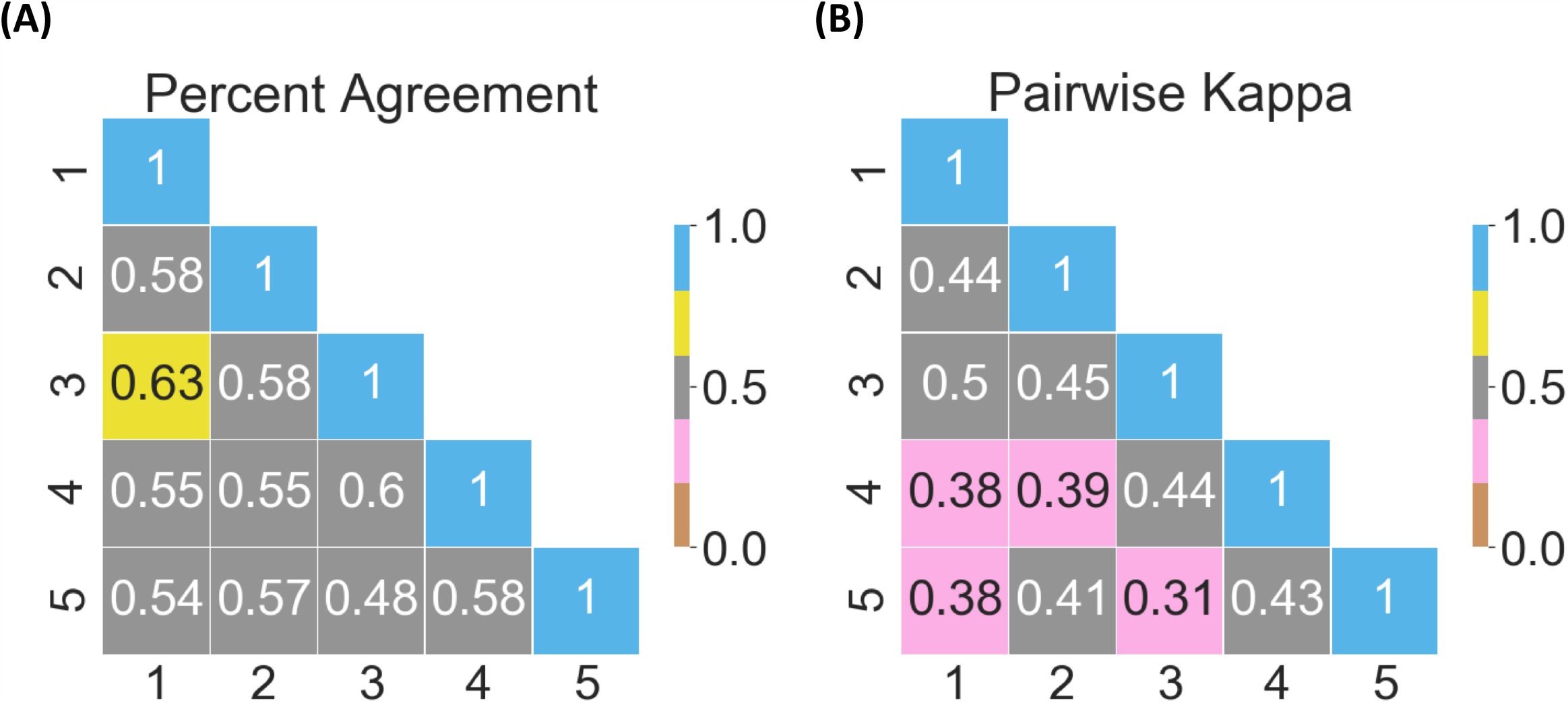
Pathologist-level IFTA grading. (A) Pairwise values of percentage agreement between the nephropathologists are shown on the cases obtained from The Ohio State University Wexner Medical Center (OSUWMC). The values were normalized to lie between 0 and 1. (B) Pairwise kappa scores between the nephropathologists on the OSUWMC data are shown. The kappa values range from 0 to 1, where 0 indicates no agreement and 1 indicates perfect agreement.

The DL model (glpathnet) accurately predicted the IFTA grade on the OSUWMC data (Accuracy=71.8±5.3%), based on 5-fold cross validation (**Figure 4**). The patch-level model predictions also consistently predicted IFTA grade, as indicated by the class-level receiver operating characteristic (ROC) curves (**Figures 4A-4D**). For the “minimal” IFTA label, the patch-level cross-validated model resulted in area under ROC curve (AUC) of 0.65±0.04. For the “mild” IFTA label, the AUC was 0.67±0.04, for the “moderate” IFTA label, the AUC was 0.68±0.09 and for the “severe” IFTA label, the AUC was 0.76±0.06. Also, for each class label, the cross-validated model performance on the WSIs was evaluated by computing mean precision, mean sensitivity and mean specificity along with their respective standard deviations. For the “minimal” IFTA label, the precision was 0.82±0.17, the sensitivity was 0.77±0.08, and the specificity was 0.93±0.07. For the “mild” IFTA label, the precision was 0.71±0.15, the sensitivity was 0.68±0.10, and specificity was 0.91±0.05. For the “moderate” IFTA label, the precision was 0.82±0.10, the sensitivity was 0.73±0.20, and specificity was 0.93±0.05. Finally, for the “severe” IFTA label, the precision was 0.65±0.06, the sensitivity was 0.72±0.16, and specificity was 0.88±0.04. It must be noted that due to the nature by which specificity was computed on the model (i.e. “minimal” vs “not-minimal”, “mild” vs “not-mild”, “moderate” vs “not-moderate” and “severe” vs “not-severe”), the values were generally higher than precision and sensitivity for all cases.

**Figure 4.**
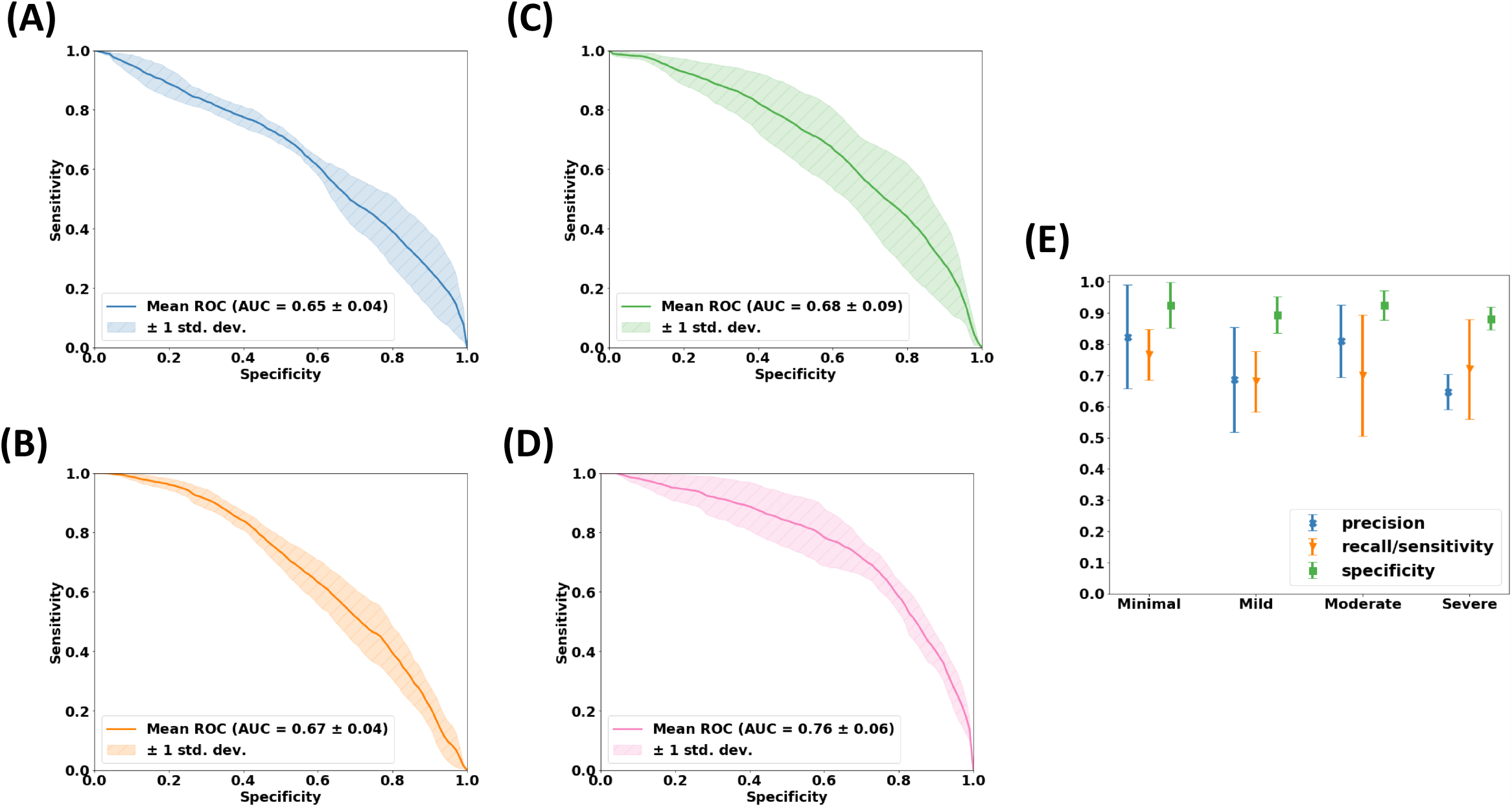
Deep learning model performance on The Ohio State University Wexner Medical Center dataset. (A-D) Patch-level performance of the 5-fold cross validated model is shown for each IFTA grade. In (A), the receiver operating characteristic (ROC) curve for the “minimal” grade is shown whereas in (B), the ROC curve for the “mild” grade is shown. In (C) and (D), the ROC curves for “moderate” and “severe” grades are shown. (E) Model performance including precision, sensitivity and specificity on the entire WSIs is shown for each IFTA grade.

Based on 5-fold cross validation, we found that there was good agreement between the true and predicted IFTA grades on the OSUWMC data, (*κ*=0.62±0.07) (**Figure S3**). Class activation mapping (CAM) was performed on the WSIs to explore the regions that are highly associated with the output class label (**Figures 5 & S4**). Two different strategies were used to generate CAMs. The first method generated CAMs at the WSI (or global) level without utilizing the local features, while the other method generated CAMs that synthesized features at the local and global level. We observed that although both these strategies generated CAMs that highlighted image sub-regions, CAMs based on the model that combined local and global representations showed higher association with the output label of interest. We also generated patch-level probabilities that had high degree association with the IFTA grade (**Figure 6**). Each image patch and its set of probability values were reviewed by the nephropathologist (C.A.C.), who confirmed that patch-level patterns were consistent with model predictions of the corresponding IFTA grades. Note that C.A.C. reviewed the patch-based results after she performed IFTA grading on all the WSIs, so she did not get biased by the model results during IFTA grading. We must note that the IFTA grade was based on WSI-level estimation and the probabilities were predicted at patch-level.

**Figure 5.**
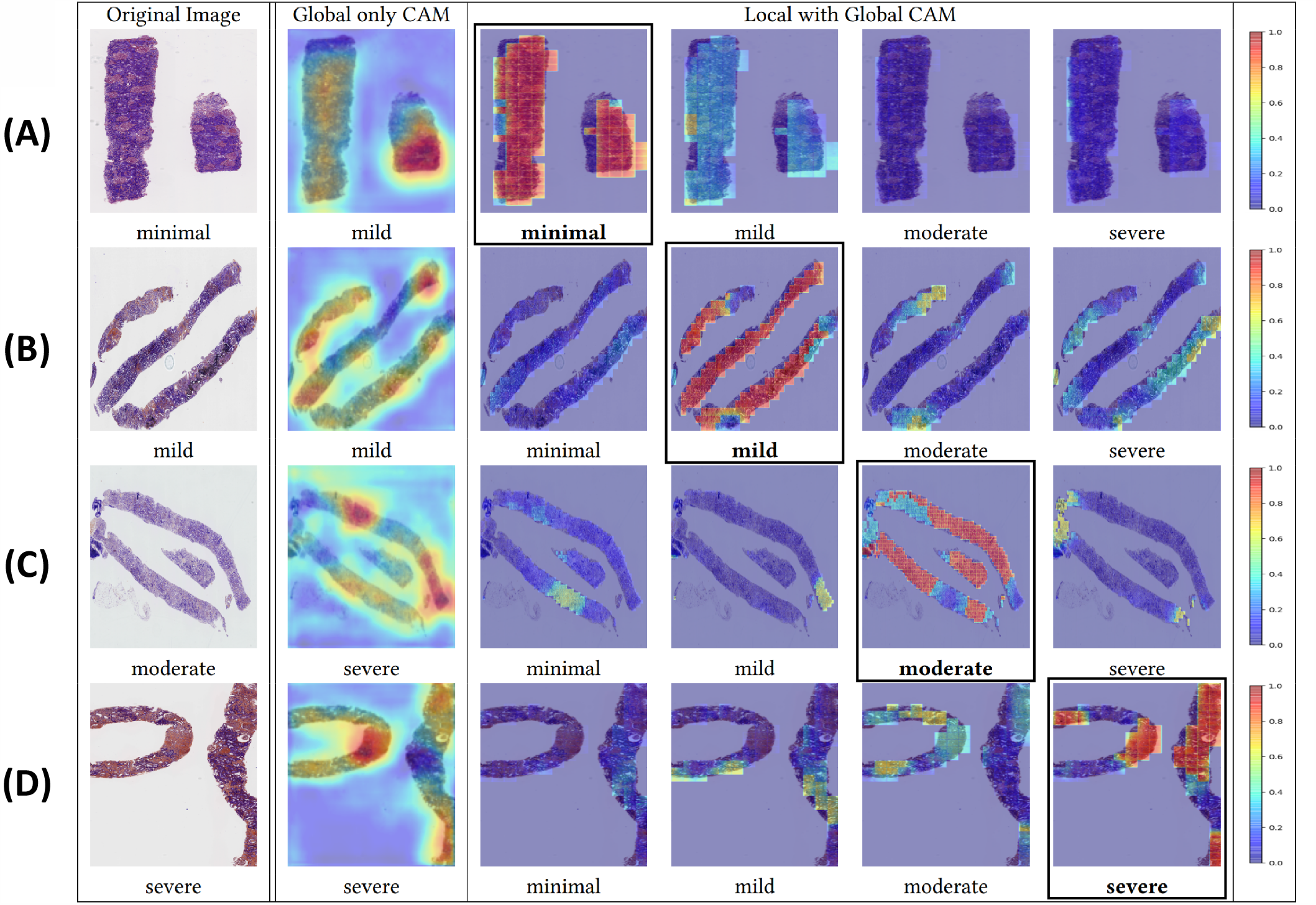
Visualization of discriminatory regions within the pathology images. The first column represents the original whole slide images (WSIs) along with the ground truth labels derived using majority voting on the pathologists’ IFTA grades. The second column shows global class activation maps (CAMs) generated on the entire WSI and the global CAM-based model predictions. The third to sixth columns show CAMs derived by combining local and global representations for each class label along with their corresponding model predictions. In the first row (A), a case with a ‘minimal’ IFTA grade is shown. The approach that used global CAMs only predicted the IFTA grade as ‘mild’, whereas the approach using local and global CAMs correctly predicted the IFTA grade as ‘minimal’. In the second row (B), a case with a ‘mild’ IFTA grade is shown. Both the approaches that used global CAMs only and the one that used local and global CAMs correctly predicted the IFTA grade as ‘mild’. In the third row (C), a case with a ‘moderate’ IFTA grade is shown. The approach that used global CAMs only predicted the IFTA grade as ‘severe’, whereas the approach using local and global CAMs correctly predicted the IFTA grade as ‘moderate’. In the fourth row (D), a case with a ‘severe’ IFTA grade is shown. Both the approaches that used global CAMs only and the one that used local and global CAMs correctly predicted the IFTA grade as ‘severe’. All these cases were obtained from the Ohio State University Wexner Medical Center.

**Figure 6.**
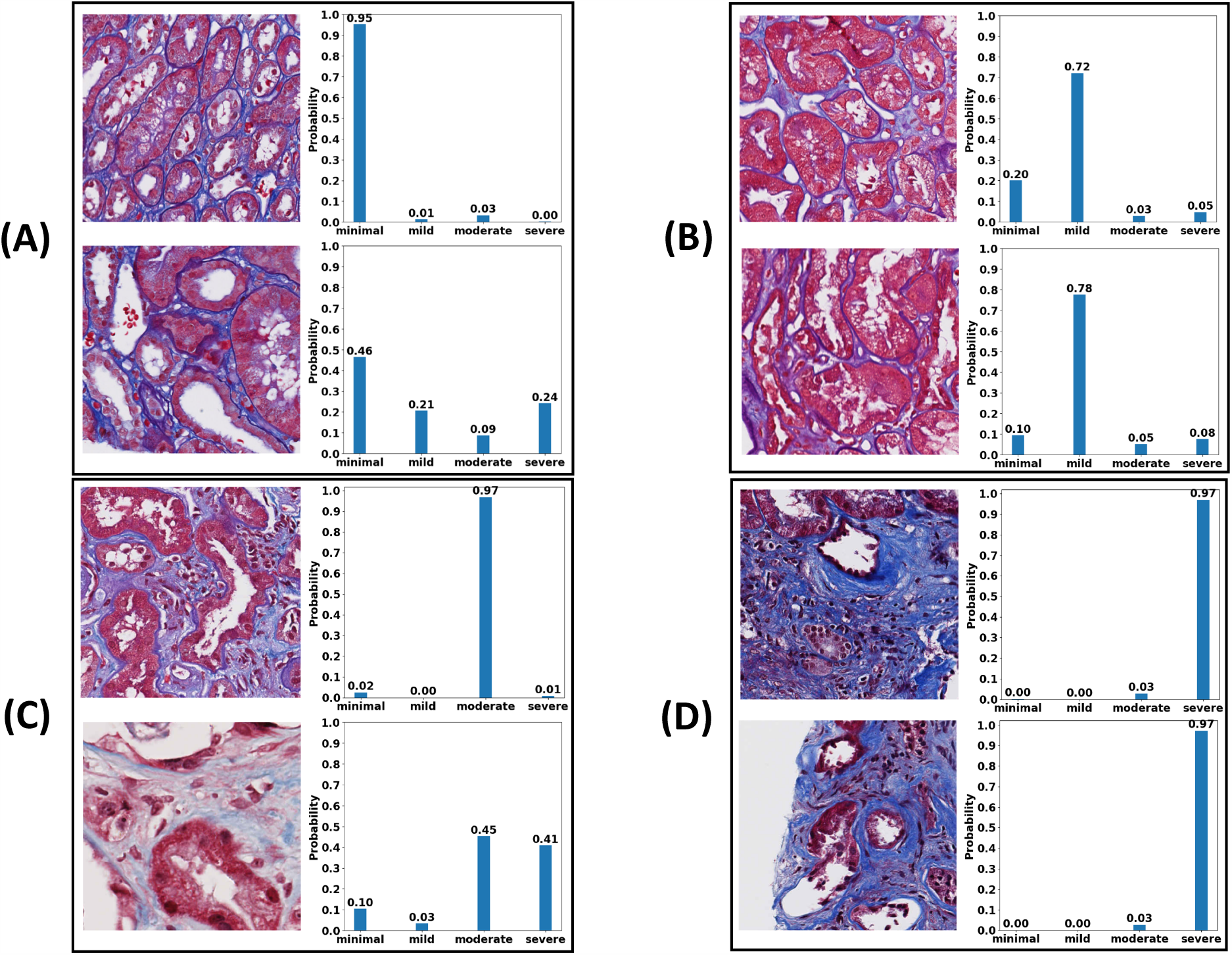
Patch-level probabilities of the deep learning model. Selected image patches and their corresponding probability values for each IFTA grade are shown. The set of image patches in (A) shows the ones with “minimal” IFTA, the patches in (B) indicate the ones with “mild” IFTA, the cases in (C) show image patches with “moderate” IFTA and finally, the image patches in (D) indicate the cases with “severe” IFTA. All the images patches and their corresponding probability values were reviewed by a nephropathologist (C.A.C.).

While the 5-fold cross-validated model on the OSUWMC dataset generated convincing results, we also wanted to understand how glpathnet performed on an external dataset. The cross-validated model was used to predict IFTA grade on the KPMP data. The entire process including random data split followed by model training using OSUWMC data and prediction on KPMP data was repeated 5 times, and average model performance was reported (Accuracy=65.0±4.2%) (**Figure 7A**). Also, for each class label, the cross-validated model performance on the WSIs was evaluated by computing mean precision, mean sensitivity and mean specificity along with their respective standard deviations. For the “minimal” IFTA label, the precision was 0.82±0.06, the sensitivity was 0.73±0.08, and the specificity was 0.78±0.08. For the “mild” IFTA label, the precision was 0.55±0.08, the sensitivity was 0.57±0.17, and specificity was 0.87±0.04. For the “moderate” IFTA label, the precision was 0.64±0.05, the sensitivity was 0.53±0.19, and specificity was 0.92±0.03. We did not compute the performance scores for the “severe” IFTA label because none of the cases from the KPMP dataset were graded as “severe” IFTA. Even for those cases, we observed that CAMs based on the model that combined local and global representations had a better association with the output label of interest (**Figures 7B-7D**). Model performance between these cohorts featuring broad variance in slide staining protocols, slide digitization, geographic location and recruitment criteria suggests a good degree of generalizability. Of note, the DL model outperformed the traditional machine learning model (WND-CHARM), which was constructed using generic image descriptors from the WSIs (**Table 2**). Specifically, WND-CHARM achieved an accuracy of 21.4±7.5% on the OSUWMC dataset and an accuracy of 35.0±14.7% on the KPMP dataset. These results underscore the advantage of utilizing DL for predicting IFTA grade on digitized kidney biopsies.

**Table 2.** Performance of the traditional machine learning model. A machine learning model based on WND-CHARM was constructed by deriving ∼3,000 features from the whole slide image data obtained from the Ohio State University Wexner Medical Center (OSUWMC) to predict IFTA grade. The trained model was then used to predict on the data obtained from the Kidney Precision Medicine Project (KPMP). (A) Performance of the model after 5-fold cross validation on the OSUWMC dataset is shown. (B) Performance of the trained model using OSUWMC dataset and predicted on the KPMP dataset is shown.

| Description | Minimal | Mild | Moderate | Severe |
| --- | --- | --- | --- | --- |
| Precision | 0.12±0.15 | 0.25±0.13 | 0.22±0.12 | 0.29±0.18 |
| Recall/Sensitivity | 0.10±0.13 | 0.40±0.17 | 0.27±0.13 | 0.23±0.17 |
| Specificity | 0.89±0.09 | 0.59±0.14 | 0.67±0.12 | 0.86±0.07 |

(B)
| Description | Minimal | Mild | Moderate | Severe |
| --- | --- | --- | --- | --- |
| Precision | 0.52±0.26 | 0.30±0.40 | 0.26±0.20 | N/A |
| Recall/Sensitivity | 0.49±0.30 | 0.07±0.08 | 0.27±0.20 | N/A |
| Specificity | 0.63±0.24 | 0.88±0.17 | 0.79±0.23 | N/A |

**Figure 7.**
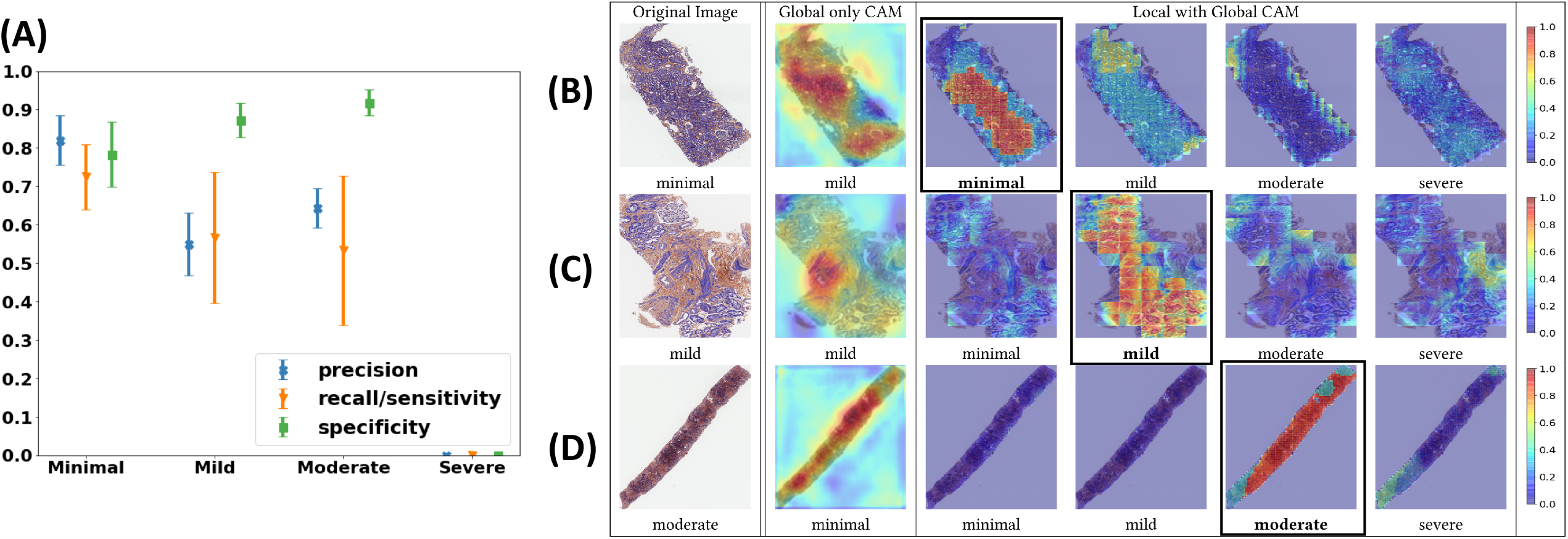
Deep learning model performance on the Kidney Precision Medicine Project dataset. (A) Model performance including precision, sensitivity and specificity on the entire whole slide images is shown for each IFTA grade (n=28). Note that performance scores for the “severe” IFTA label were not computed because none of the cases were graded as “severe” IFTA. (B-D) Class activation maps (CAMs) were generated on the dataset. The first column represents the original whole slide images (WSIs) along with the ground truth labels derived using majority voting on the pathologists’ IFTA grades. The second column shows global CAMs generated on the entire WSI and the global CAM-based model predictions. The third to sixth columns show CAMs derived by combining local and global representations for each class label along with their corresponding model predictions. In the first row (B), a case with a ‘minimal’ IFTA grade is shown. The approach that used global CAMs only predicted the IFTA grade as ‘mild’, whereas the approach using local and global CAMs correctly predicted the IFTA grade as ‘minimal’. In the second row (C), a case with a ‘mild’ IFTA grade is shown. Both the approaches that used global CAMs only and the one that used local and global CAMs correctly predicted the IFTA grade as ‘mild’. In the third row (D), a case with a ‘moderate’ IFTA grade is shown. The approach that used global CAMs only predicted the IFTA grade as ‘minimal’, whereas the approach using local and global CAMs correctly predicted the IFTA grade as ‘moderate’.

## Discussion

As kidney disease remains to be the 9^th^ leading cause of death in the US (31), current options for clinical management are too expensive and fail to provide a high quality of life. The recent initiative by the White House known as *Advancing American Kidney Health* is a clear indication that preventing kidney failure through better diagnosis, treatment and preventative care is among the nation’s top healthcare priorities (32). The complexity of clinical management is compounded by the fact that methods to grade disease are manual and driven by visual estimation, leading to variability among the expert pathologists. In presence of these challenges, the NIH along with other stakeholders have recently launched nationwide initiatives (KPMP (https://www.kpmp.org) & KidneyX (https://www.kidneyx.org)) to tackle global kidney disease burden. These projects are expected to provide tremendous opportunities to accelerate innovation, re-define kidney disease in molecular terms and identify novel targeted therapies, as well as transform kidney care and management. Such goals can be achieved when rich patient data can be analyzed using state-of-the-art technologies, visualized with the help of next-generation software tools and shared openly in the public domain. Motivated by such developments and in anticipation of the enormous amount of data that is planned to be generated, we developed a data analytics framework that can analyze digitized kidney biopsies at the level of an expert pathologist. Specifically, we selected automated IFTA grading as our task because fibrosis on kidney biopsy is a known structural correlate of progressive and chronic kidney disease (20, 33).

Despite knowing the putative link between IFTA grade and disease prognosis, there remains uncertainty about how to best measure fibrosis within the kidney. Farris & Alpers provided an interesting perspective on this aspect in their recent article (2), where they argue that current analytic approaches generally avoid rigorous assessment of various aspects related to characterizing fibrosis of the tubulointerstitium. They note that human reproducibility is not generally high because there is no agreeable definition on how to measure IFTA. For example, some consider percent interstitial fibrosis to be percent of overall tissue occupied by fibrous tissue, whereas others note that the percent of fibrosis to be the percentage of tissue that is abnormal. In clinical practice however, people often refer to scoring systems defined by established national and international working groups (Renal Pathology Society Working Group, Banff classification, etc.) for grading fibrosis. Morphometric analysis can also be performed to evaluate renal fibrosis as this approach can bring efficiency, reproducibility and functional correlation (3). These developments lend themselves to using more advanced computer methods such as machine learning on digitized images for IFTA grading.

The novelty of glpathnet is underscored by the fact that it combines local representations to quantify features at the image patch level as well as at the WSI-level to accurately predict the IFTA grade. A combination of both these assessments provides a comprehensive evaluation of IFTA. This methodology appears to capture the typical workflow of pathologists as they examine the WSIs by performing manual operations such as panning across the WSI to perceive the overall context, zooming in and out of specific WSI regions to evaluate the local pathology, and finally combining the information learned from both these steps to determine the IFTA grade. We must however acknowledge that the nephropathologist’s clinical impression and diagnosis is based on contextual factors above and beyond visual inspection of a lesion in isolation.Nevertheless, by identifying WSI regions using CAMs that are highly indicative of a class label, our approach provides a quantitative basis by which to interpret the model-based predictions rather than viewing DL methods as black-box approaches. As such, our approach stands in contrast to other methods that rely on expert-driven annotations and segmentation algorithms that attempt to quantify histological regions and derive information for pathologic assessment (12, 15-18).

Saliency mapping based on CAMs is increasingly being considered as a framework to generate visual interpretations of model predictions by highlighting image sub-regions that are presumably correlated with the outputs of interest (34-43). These ‘heat maps’ can be generated for any input image that is associated with an output class label (i.e., IFTA grade). The underlying assumption is that the heat map representation highlights pixels of the image that triggers the model to associate the image with a particular class label. Our DL strategy that combined local and global representations consistently predicted the correct IFTA grade and generated interpretable CAMs. This strategy turned out to be superior than using only the global representations, where several image sub-regions where the CAMs were highlighted appear to have high correlation with the output class label.

This study has a few limitations. We relied only on WSIs derived using the trichrome stain, as this is commonly used by nephropathologists. Past studies have found that trichrome-stained slides can produce cost-effective, efficient, reproducible and functional correlations with outcomes such as eGFR (2, 3); some pathologists, however, rely on other protocols such as Hematoxylin & Eosin, Periodic acid–Schiff, Jones Methenamine Silver or Sirius Red staining to grade fibrosis (2, 44-46). Our DL framework has the potential to be applied effectively to WSIs generated with other staining protocols. We partially demonstrated a solution to this issue because the OSUWMC cases that we obtained included both in-house cases and consults from external institutions, indicating that our model performed well on cases that may have employed different staining techniques. Furthermore, we recognize that our DL framework provides an automated approach to IFTA grading to assist the pathologist rather than replacing the human factor. Nevertheless, the ability to classify WSIs using a computer with the accuracy of an experienced nephropathologist has the potential to inform pathology practices, especially in resource-limited settings.

In conclusion, we demonstrated the effectiveness of capturing localized morphological along with WSI-level contextual features using an advanced DL architecture (glpathnet) to mimic the pathologist’s approach to IFTA grading. It is possible to use glpathnet to study other organ-specific pathologies focused on evaluating fibrosis, as well as WSIs generated using other histological staining protocols. Our proposed framework to local and contextual IFTA grading can serve as an ‘analysis template’ for researchers and practitioners when new data from cohort studies such as KPMP become available. Further validation of the glpathnet across different pathology practices and patient populations is necessary to study if it can be effective across the full distribution and spectrum of fibrotic lesions.

## Acknowledgments

We thank Ms. Ashveena Dighe, Dr. Robyn McClelland and Dr. Jeffrey Hodgin for providing access to the KPMP data. The KPMP is funded by the following grants from the NIDDK: U2C DK114886, UH3DK114861, UH3DK114866, UH3DK114870, UH3DK114908, UH3DK114915, UH3DK114926, UH3DK114907, UH3DK114920, UH3DK114923, UH3DK114933, and UH3DK114937.

## Funding

VBK is supported by grants from the National Center for Advancing Translational Sciences, National Institutes of Health (NIH), through BU-CTSI Grant (1UL1TR001430), a Scientist Development Grant (17SDG33670323) and a Strategically Focused Research Network (SFRN) Center Grant (20SFRN35460031) from the American Heart Association, and a Hariri Research Award from the Hariri Institute for Computing and Computational Science & Engineering at Boston University, and NIH grants (R01-AG062109 and R21-CA253498). VCC is supported by NIH grants (R21-DK119740 and R01-HL132325). PB is supported by the German Research Foundation (DFG; SFB/TRR57, SFB/TRR219, BO3755/3-1, BO3755/9-1, BO3755/13-1), the German Federal Ministries of Education and Research (BMBF: STOP-FSGS-01GM1901A), Health (DEEP LIVER, ZMVI1-2520DAT111) and Economic Affairs and Energy (EMPAIA). SSW is supported by NIH grants (R21-DK119751 and U01-DK085660). MB is supported by National Science Foundation grants (#1838193 and #1551572).

## Disclaimer

The content is solely the responsibility of the authors and does not necessarily represent the official views of the National Institutes of Health.

## Supplementary figures and tables

**Table S1.**
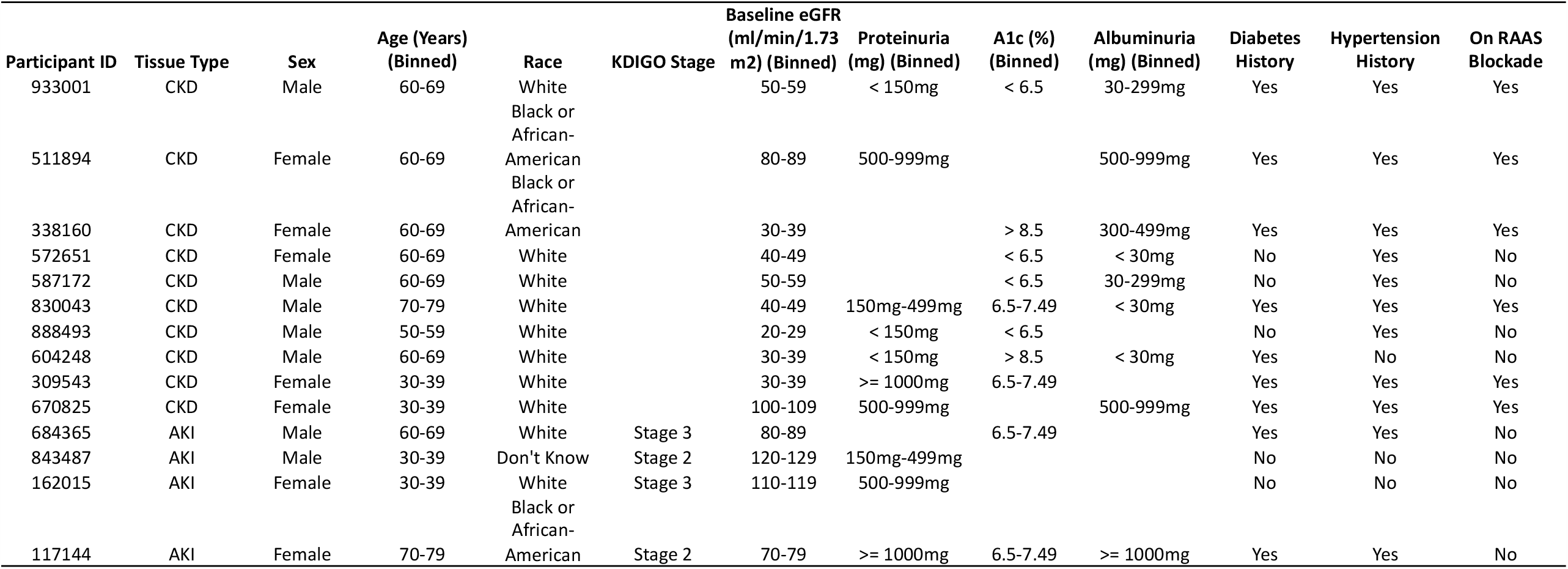
Kidney Precision Medicine Project cohort. Clinical and demographic characteristics of the KPMP participants (n=14) are shown. For each participant, two trichrome stained whole slide images in SVS format were available. More info on the KPMP project is available at https://www.kpmp.org.

**Figure S1.**
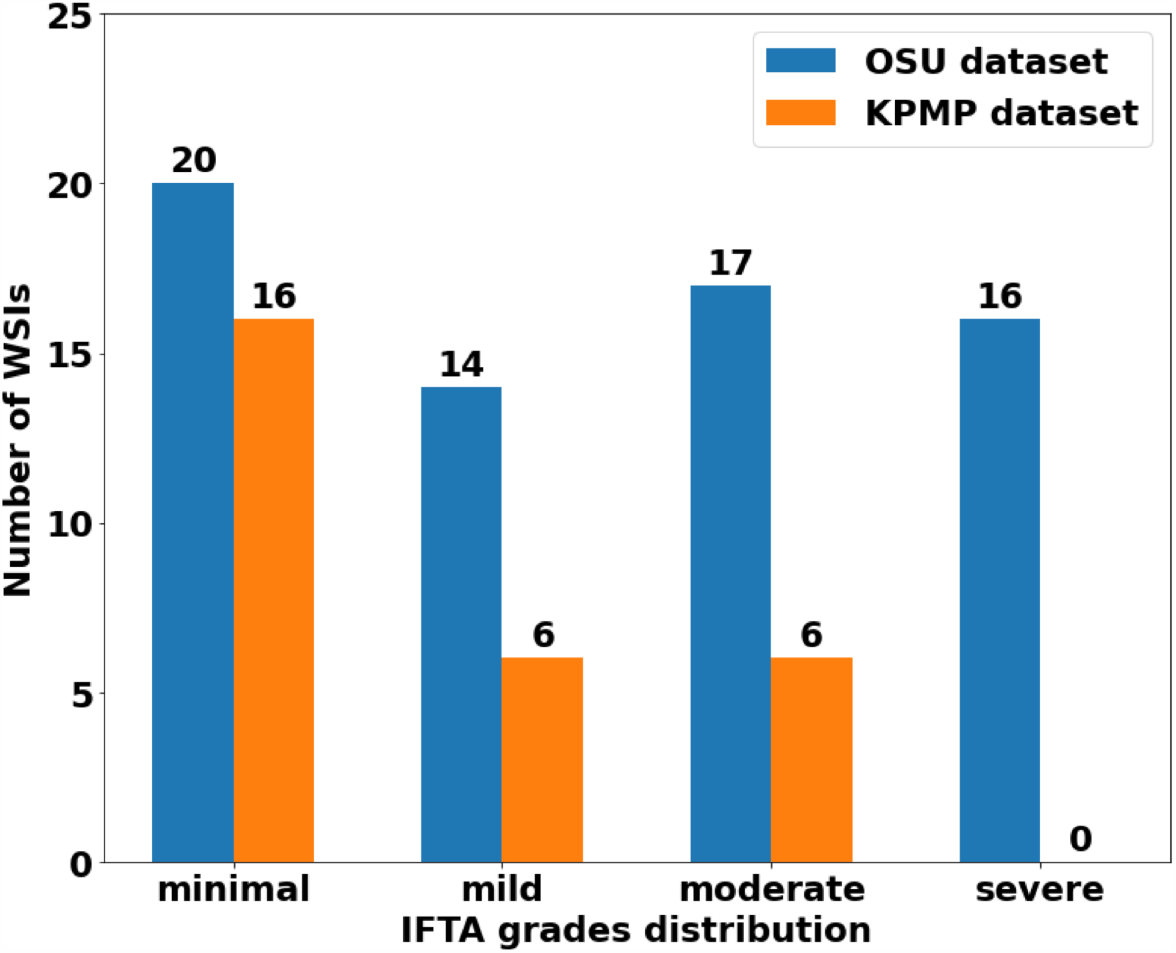
WSI distribution. Number of WSIs per IFTA grade in the OSUWMC and KPMP datasets are shown.

**Figure S2.**
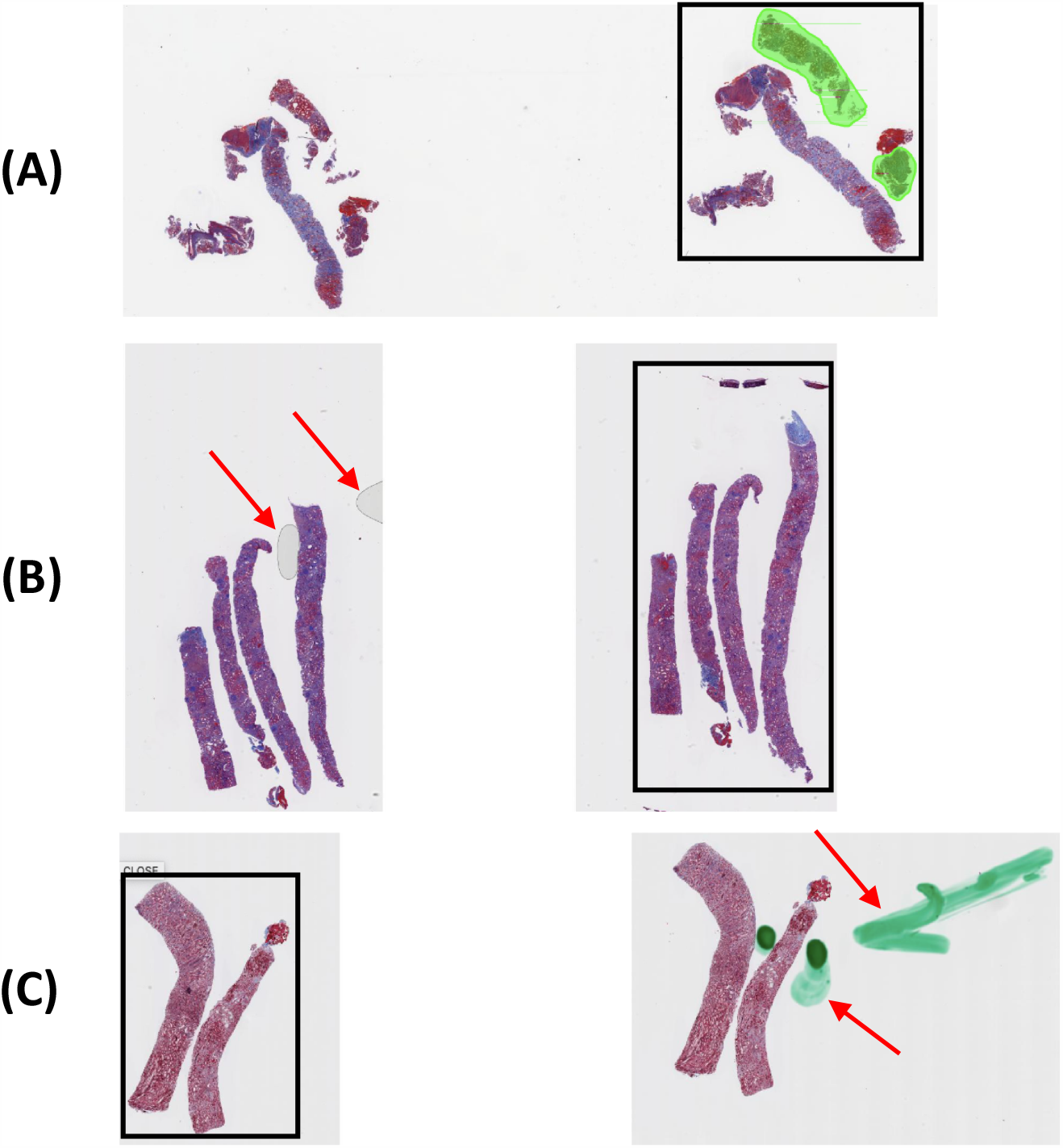
Quality check and ROI selection. A manual quality check was performed by a nephropathologist to identify and select whole slide image regions that served as inputs to the DL model. (A) This case shows that the core on the extreme right containing both the cortex and the medulla was selected for further analysis. (B) This case shows the presence of air bubbles (indicated by red arrows) on the second core. The third core was selected for further analysis. (C) This case shows staining artifacts (indicated by red arrows) on the second core. The first core was selected for further analysis.

**Table S2.**
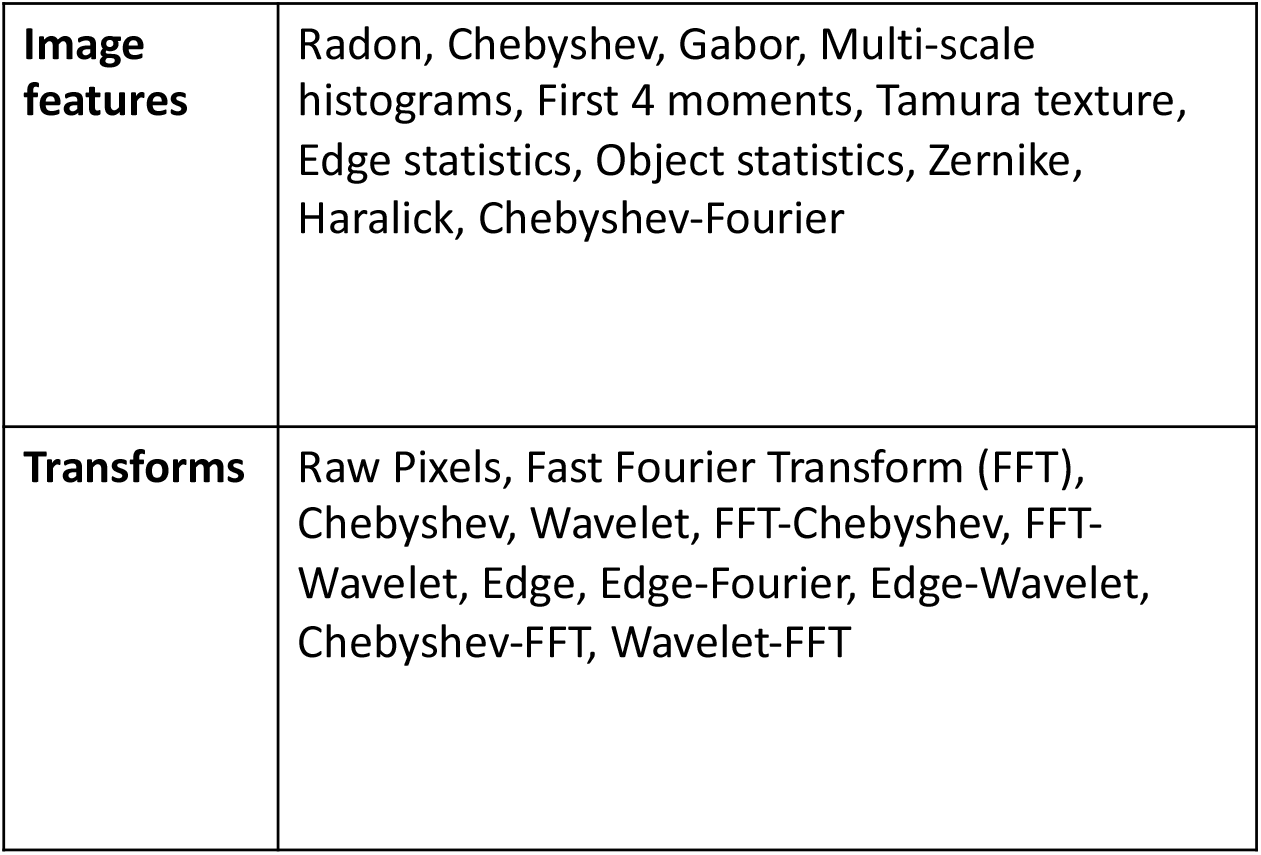
WND-CHARM features. The table shows the image features and the transforms used in WND-CHARM, which is based on traditional and classic image processing. More details can be found in the original paper (29).

**Figure S3.**
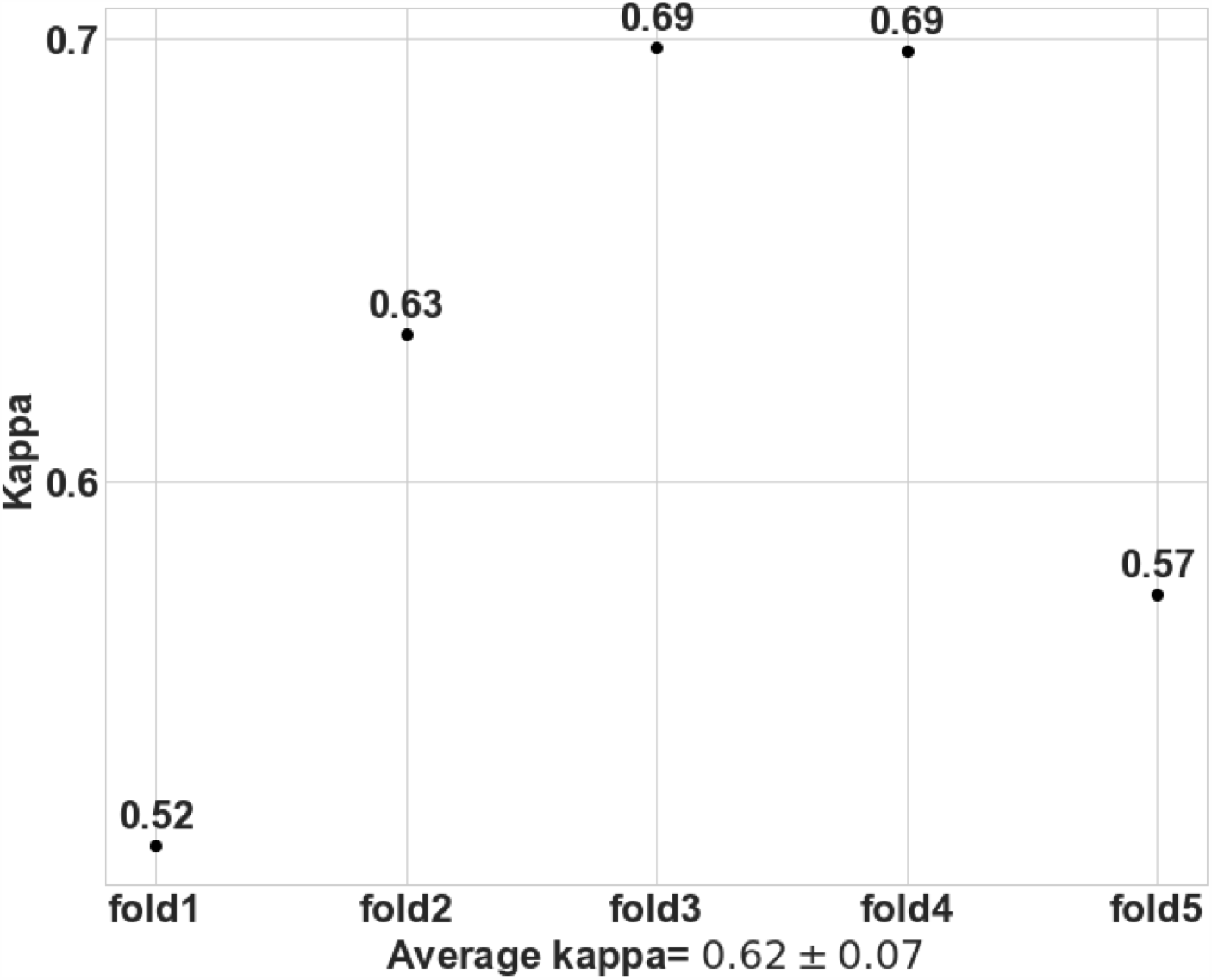
Model agreement with the ground truth. We performed 5-fold cross validation on the OSUWMC dataset to evaluate the DL model. For each fold, Kappa was computed to evaluate the agreement of the model with the ground truth defined using majority voting on the IFTA grading performed by the nephropathologists.

**Figure S4.**
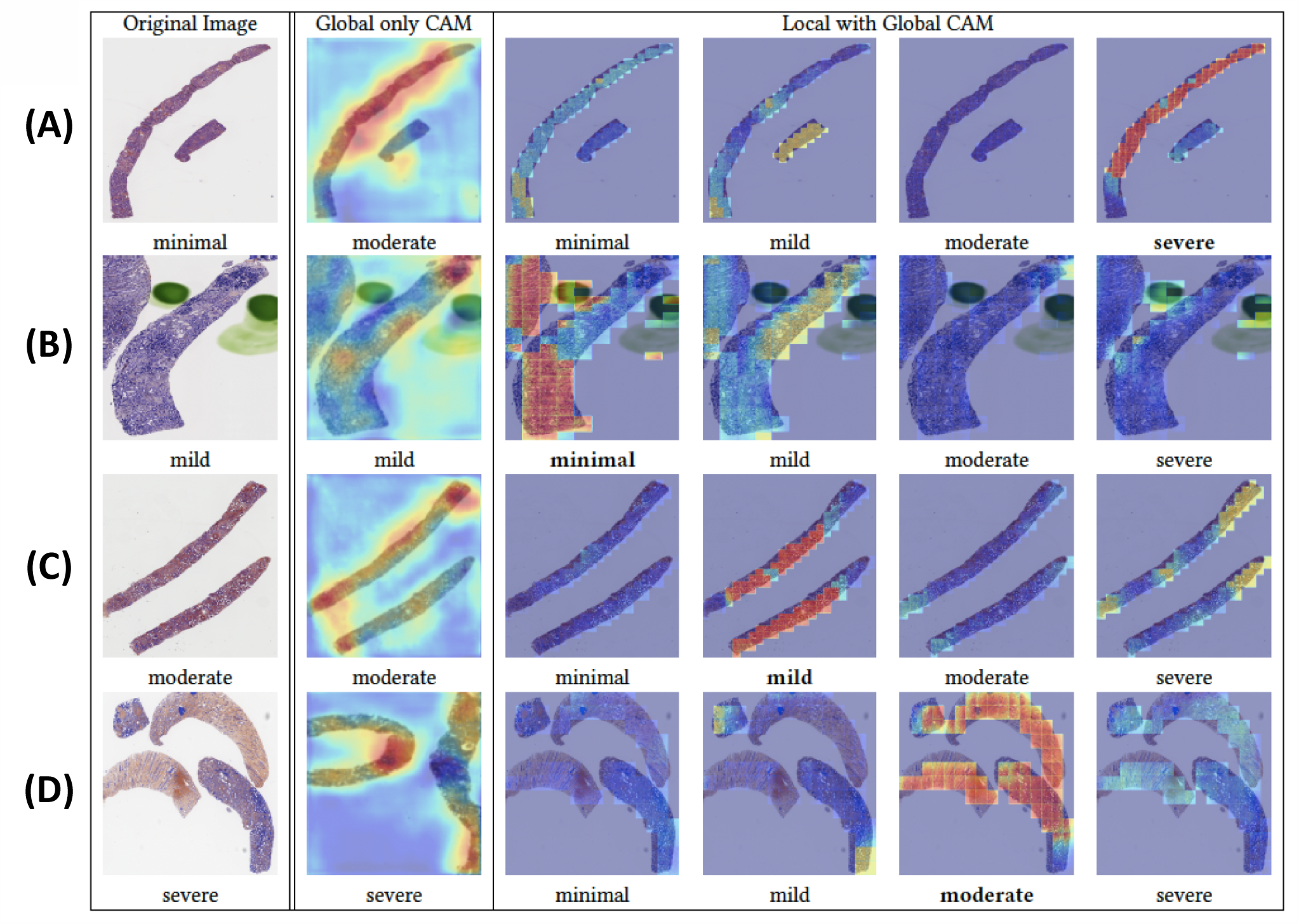
Visualization of discriminatory regions within the pathology images. The first column represents the original whole slide images (WSIs) along with the ground truth labels derived using majority voting on the pathologists’ IFTA grades. The second column shows global class activation maps (CAMs) generated on the entire WSI and the global CAM-based model predictions. The third to sixth columns show CAMs derived by combining local and global representations for each class label along with their corresponding model predictions. In the first row (A), a case with a ‘minimal’ IFTA grade is shown. The approach that used global CAMs only predicted the IFTA grade as ‘moderate’, whereas the approach using local and global CAMs predicted the IFTA grade as ‘severe’. In the second row (B), a case with a ‘mild’ IFTA grade is shown. In this case, the global only CAM-based model correctly predicted the IFTA grade, whereas the local and global CAM-based approach predicted the IFTA grade as ‘minimal’. In the third row (C), a case with a ‘moderate’ IFTA grade is shown. The approach that used global CAMs only correctly predicted the IFTA grade as ‘moderate’, whereas the approach using local and global CAMs incorrectly predicted the IFTA grade as ‘mild’. In the fourth row (D), a case with a ‘severe’ IFTA grade is shown. The approach that used global CAMs only correctly predicted the IFTA grade as ‘severe’, whereas the approach using local and global CAMs correctly predicted the IFTA grade as ‘moderate’. All these cases were obtained from the Ohio State University Wexner Medical Center.

## Notes

### Competing Interest Statement

The authors have declared no competing interest.

### Author Declarations

Renal biopsy as well as patient data collection, staining and digitization followed protocols approved by the Institutional Review Board at OSUWMC (Study number: 2018H0495)

